# Distinct Patterns of Mobility Recovery After Stroke Using Routine Clinical Data

**DOI:** 10.64898/2026.07.08.26357600

**Authors:** Margaret A. French, Elizabeth B. Marsh, Ryan Roemmich, Preeti Raghavan

## Abstract

**Background:** Mobility recovery after stroke is highly variable, yet is typically described using average patterns that obscure meaningful differences between individuals. Identifying distinct recovery trajectories may improve prognostication and guide rehabilitation strategies.

**Methods:** We conducted a retrospective cohort study of adults admitted for stroke to a large health system between 2016 and 2024. Mobility was assessed using Activity Measure for Post-Acute Care (AM-PAC) Basic Mobility, which was collected during routine clinical care. Growth mixture modeling was used to identify subgroups with distinct mobility recovery trajectories during the first 180 days after stroke. Subgroups were then characterized with baseline personal and clinical characteristics.

**Results:** Seven hundred and fifty individuals contributed 3,389 mobility observations (median 4 per person). A five-class solution was selected based on model fit and classification quality. Distinct trajectories were identified: low stable (n=127), low rapidly improving (n=29), mid declining (n=169), mid improving (n=365), and high stable (n=60). Subgroups differed in both baseline mobility and patterns of change over time, with some demonstrating improvement, others remaining stable, and one declining. Individuals in improving subgroups were generally younger, more likely to be independent before stroke, received physical therapy on a greater proportion of hospital days, and were more frequently discharged to inpatient rehabilitation. In contrast, those in low or declining trajectories had lower baseline function, longer hospital stays, and were more likely to be discharged to skilled nursing facilities.

**Conclusions:** The distinct mobility recovery trajectories identified in this work reflect the heterogeneity present in routine clinical practice. Subgroups differed in both recovery patterns and characteristics. Early identification of trajectory membership may improve prognostication and inform more targeted rehabilitation strategies.

## Introduction

Many individuals with stroke have mobility deficits that persist and contribute to long-term disability.^1,2^ These mobility challenges include difficulties with changing positions and walking and are associated with negative health outcomes, such as hospital readmissions, recurrent stroke, falls, and reduced quality of life.^3–8^ Improving mobility is also consistently identified as a primary goal of individuals with stroke,^9–11^ making mobility recovery a primary target for post-stroke rehabilitation. Understanding the characteristics of individuals that influence distinct patterns of recovery after stroke is critical for informing patient expectations, guiding clinical decision-making, and selecting appropriate rehabilitation interventions. Without this understanding, clinicians rely on average recovery patterns that obscure meaningful differences between patients, contributing to inaccurate expectations and inappropriate rehabilitation strategies. However, such an understanding remains limited, in part due to substantial heterogeneity in mobility recovery that has not been well characterized.

Understanding variability in recovery requires examining trajectories that reflect both the rate and extent of recovery over time. To date, mobility recovery has most often been summarized using group averages of change,^12–14^ hiding differences between individuals. Recent work related to upper extremity and global recovery after stroke has begun to address this limitation by identifying subgroups with distinct recovery trajectories defined by the rate and extent of recovery.^15–17^ Similar trajectory-based approaches have been used in other patient groups, including low back pain^18,19^ and total knee arthroplasty,^20,21^ to better characterize heterogeneity in functional outcomes. However, this approach has not been applied to post-stroke mobility, leaving the field with limited understanding of distinct recovery patterns following stroke.

Trajectory-based approaches require large samples that are rarely obtained in post-stroke rehabilitation trials. In previous work in post-stroke upper extremity recovery, data from multiple clinical trials were combined to obtain a large cohort. While this is one option, clinical trial populations often include selected groups that do not fully capture the diversity of patients seen in routine clinical practice.^22,23^ Electronic health record (EHR) data offer an alternative data source to characterize recovery trajectories from large, real-world populations, particularly when consistent outcomes are collected across healthcare settings.^24,25^ With improving consensus on how to measure function after stroke, EHRs are increasingly suitable for this type of analysis, allowing data collected during clinical care to identify recovery trajectories that reflect the heterogeneity observed in practice. Leveraging these data may improve prognostication and inform stratification of patients for more targeted rehabilitation interventions and future clinical trials.

Therefore, our purpose was to 1) identify subgroups of individuals based on mobility recovery during the first six months (180 days) after stroke using longitudinal mobility data captured during routine clinical care and 2) characterize those subgroups based on baseline personal (e.g., age, sex, previous level of function) and clinical characteristics (e.g., length of stay, initial functional status, discharge location). To address this objective, we applied growth mixture modeling to longitudinal mobility data derived from the EHR, with mobility operationalized using the Activity Measure for Post-Acute Care (AM-PAC) Basic Mobility score.

## Methods

### Participants

In this retrospective cohort study, we included adults admitted with stroke to two hospitals within Johns Hopkins Medicine (Johns Hopkins Hospital or Johns Hopkins Bayview) between July 1, 2016, and June 30, 2024. Stroke was identified by ICD-10 codes within the EHR (Supplemental Methods). To be included, individuals needed to be discharged alive and have three measurements of functional mobility during the first 180 days after stroke, with date of admission serving as a proxy for time of stroke. Because the objective was to identify mobility recovery trajectories, analyses were restricted to individuals with sufficient longitudinal mobility assessments to characterize change over time within routine clinical care. We required that the first measurement occur during the hospital admission, and the last measurement occur between 91- and 180-days post-stroke, with the second one anytime between the two. Individuals were excluded if they had a hospital stay greater than 50 days or were discharged to another hospital, psychiatric hospital, or other unusual location (e.g., prison). The Johns Hopkins Hospital IRB approved this study (IRB00291279) and granted a waiver of consent.

### Measurement of Physical Function

We used the AM-PAC Basic Mobility to quantify functional mobility.^26^ This patient-, proxy-, or clinician-reported measure includes questions about how much difficulty the individual has performing common mobility tasks such as walking and stair climbing. It is systematically collected across the system during routine clinical care. During acute care and inpatient rehabilitation, the AM-PAC Inpatient Mobility Short Form, commonly called the ‘6-clicks,’^27,28^ is collected by physical therapists and nursing staff. Prior studies have demonstrated good interrater reliability for the AM-PAC across physical therapists as well as between physical therapists and nurses.^27,29^ In outpatient settings, the AM-PAC Community Mobility Short Form is collected by physical therapists.^30^ Because both forms are derived from the same AM-PAC item pool and calibrated to a common underlying metric, scores can be compared across care settings and versions of the measure.^30,31^ Although we only required three measurements of AM-PAC Basic Mobility for inclusion, we included all measures collected within 180 days of stroke in the analysis.

### Personal and Clinical Characteristics

We extracted personal characteristics including demographic information, medical history information, and social determinants of health. Demographic and medical history information were collected during clinical care and extracted from the EHR. Demographic information included age, sex, race, and previous level of function (independent/not independent). Medical history information included a history of important comorbidities including hypertension, diabetes, depression, or hyperlipidemia (present/absent). These comorbidities were identified using ICD-10 codes (Supplemental Methods). Social determinants of health were obtained from the EHR and from external sources. From the EHR, we extracted living situation (with someone/alone) and insurance type (Medicare, Medicaid, private, other). We also linked patient-level zip code information with the Neighborhood Atlas to obtain Area Deprivation Index (ADI).^32,33^ The ADI is a measure of an area’s socioeconomic resources associated with the individual’s residential zip code.^32^

Clinical characteristics were extracted from the EHR or from the hospital-specific Get-With-The-Guidelines (GWTG) database.^34^ EHR-derived variables included initial function related to activities of daily living and cognition, length of stay in the hospital, and discharge location (home, skilled nursing facility, inpatient rehabilitation facility). Initial activities of daily living (ADL) function was documented by occupational therapy using the AM-PAC Daily Activity, and initial cognitive function was documented by occupational therapy or speech-language pathology using the AM-PAC Applied Cognition. The AM-PAC Applied Cognition was not implemented as part of routine clinical care until 2018 and is therefore missing for a substantial portion of the sample. We determined whether occupational therapy or speech-language pathology were involved in the individual’s care (yes/no) via services billed during the admission. Because physical therapy most directly addresses mobility, we determined the proportion of days during the hospitalization that individuals received physical therapy. From the GWTG databases, we obtained stroke etiology (ischemic/hemorrhagic) and first NIH Stroke Scale obtained.

### Statistical analysis

To identify subgroups with distinct mobility recovery trajectories, as measured by the AM-PAC Basic Mobility, we conducted growth mixture modelling (GMM) using the lcmm (v0.9.9) package^35^ in R (v4.0.5).^36^ GMM, sometimes referred to as latent class mixed-effects models, identifies unique subgroups (often called classes) of individuals who have similar longitudinal trajectories within a heterogeneous population.^37–39^ Based on established patterns of post stroke recovery, we initially specified a quadratic time term (with month as the unit of time) to capture nonlinear change in mobility over time, along with a random intercept and random linear slope to account for inter individual variability in baseline mobility and recovery rate. We did not include a random quadratic slope because most participants contributed only three repeated measurements, which would limit model identification and stability.

We first evaluated the form of time and the random-effects structure using single-class models and likelihood ratio tests (Table S1). Although random linear slope models were theoretically appropriate, they produced unstable solutions with small classes when extended to growth mixture models, suggesting that individual-level slope variability was not supported by the data. Therefore, subsequent growth mixture models included a random intercept without a random slope. We then evaluated two variance specifications: 1) a model that constrained residual variance to be equal across classes and 2) a model that allowed residual variance to vary by class. Allowing residual variances to differ improved model fit while maintaining stable and interpretable subgroups; this specification was therefore selected as the most parsimonious representation of the data.

We fit this model with two to seven classes to evaluate a plausible range of solutions. We evaluated each candidate model with fit statistics, including the Akaike Information Criterion (AIC), Bayesian Information Criterion (BIC), sample size adjusted BIC (ssBIC), and entropy. Lower AIC, BIC, and ssBIC values and higher entropy values indicate better model fit. We also conducted Vuong–Lo–Mendell–Rubin (VLMR) likelihood ratio tests to evaluate whether the inclusion of another class significantly improved model fit, with a significant p value indicating superiority of the more complex model. In addition, we examined the mean posterior probability of class assignment for each subgroup, with values greater than 0.70 indicating adequate classification certainty.^40,41^ Finally, we considered subgroup size when evaluating and selecting the model. A general guideline for identifying stable and clinically meaningful subgroups is having at least 5% of the sample in each subgroup;^40,41^ however, this threshold was not treated as a strict selection criterion. Instead, model selection balanced statistical fit, classification quality, subgroup size, and clinical relevance rather than relying on a single criterion.

To evaluate whether the differences in subgroup trajectories reflected differential observation patterns, we examined the number of observations per participant, the follow-up duration (i.e., days between the first and last observation), and the temporal distribution of measurements across subgroups to ensure comparable observation patterns. Sensitivity analyses restricting to participants with similar observation patterns were planned but were not required given the observed similarity across subgroups.

After identifying the final model, we described each subgroup using personal and clinical characteristics. Class membership was assigned based on each individual’s highest posterior probability. Differences between subgroups were evaluated using Kruskal–Wallis tests for continuous variables and chi-squared tests for categorical variables. We did not perform post-hoc comparisons due to the descriptive nature of this analysis. Because class assignments are probabilistic and include inherent uncertainty, we conducted sensitivity analyses to evaluate the robustness of these subgroup descriptions. First, we restricted the sample to individuals with a posterior probability ≥0.7 for their assigned class and repeated subgroup comparisons using Kruskal–Wallis and chi-squared tests. Second, we performed probability-weighted analyses in which individuals contributed to each subgroup proportionally to their posterior probability of class membership. Continuous variables were summarized as weighted medians and categorical variables as weighted proportions. Because probability-weighted analyses do not yield independent group assignments, p-values were not calculated.

## Results

### Subjects

We included 750 individuals in the longitudinal mobility recovery cohort from 6,366 individuals admitted with stroke. The primary reason individuals were not included was the absence of an AM-PAC Basic Mobility assessment between 90 and 180 days after stroke (Figure 1). Consequently, the analytic cohort reflects stroke survivors with longitudinal mobility data available within the health system rather than all individuals admitted with stroke. The individuals included in the analysis were significantly younger, were more likely to be black, had significantly longer lengths of stay, and were more likely to be discharged to an inpatient rehabilitation facility after their stroke compared to those who were excluded (Table S2). Among participants with available data, included and excluded individuals had similar distributions of stroke type, baseline mobility, and activities of daily living function, and several social determinants of health (Table S2). The 750 individuals contributed 3389 observations of the AM-PAC Basic Mobility (median per person 4; interquartile range [IQR] 3-5). The median timing of the first and last AM-PAC Basic Mobility measurement was 1 day (IQR 1-1) and 141 days (IQR 115-166), respectively.

**Figure 1.**
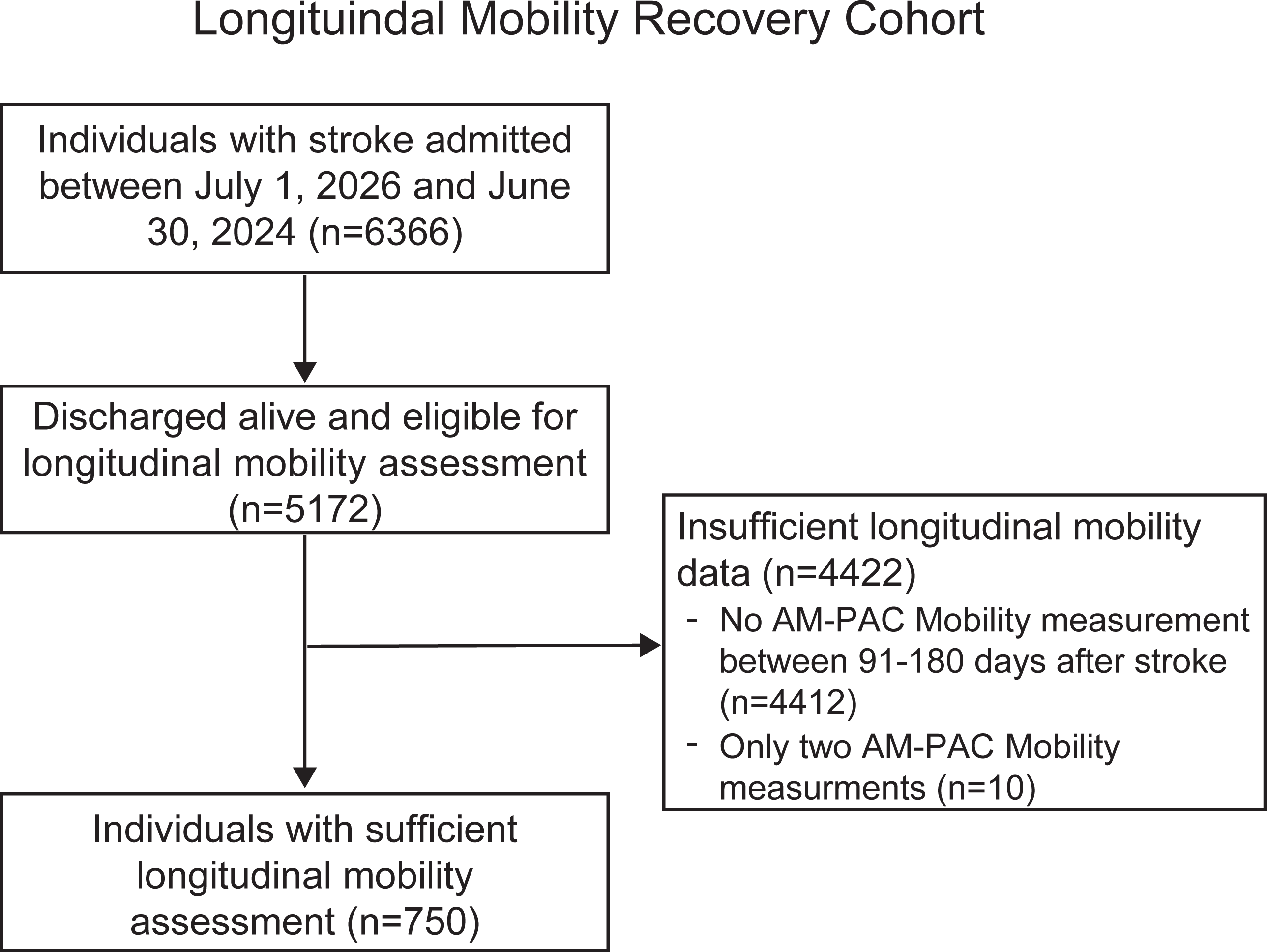
Flow diagram of cohort.

### Model selection

The five-class solution was selected based on improved model fit statistics and classification quality relative to the four-class model (Table S3). Although, information criteria (i.e., AIC, BIC, and ssBIC) continued to decrease with additional classes, models with additional classes yielded smaller subgroups and reduced classification quality. The smallest subgroup in the five-class solution included 29 individuals (3.9% of the sample), slightly below the commonly recommended 5% threshold. However, this solution demonstrated higher entropy, stronger posterior classification probabilities, and improved information criteria compared to the four-class solution. Importantly, the recovery pattern represented by this subgroup was also evident in the four-class solution, although it was combined with a larger subgroup and therefore appeared less distinct (Figure S1, class 2). We therefore selected the five-class solution because it provided greater separation of clinically interpretable recovery patterns while maintaining acceptable classification quality. The four-class solution, in which all subgroups exceeded the 5% size guideline, was evaluated as a sensitivity analysis and demonstrated similar overall recovery patterns.

### Subgroup descriptions

The five-class solution identified distinct trajectories of mobility recovery over time (Figure 2), with subgroups differing in both baseline function and rate of change (Table S4). Based on the initial functional status and the recovery trajectory, we named the four subgroups “Low stable,” “Low rapidly improving,” “Mid declining,” “Mid improving,” and “High stable.” The “Low stable” subgroup (n=127) had low baseline mobility and remained consistently low throughout follow-up, with minimal evidence of functional change. The “Low rapidly improving” subgroup (n=29) also began with low baseline mobility but demonstrated substantial improvement over time, with the steepest gains occurring early in follow-up before plateauing. The “Mid declining” subgroup (n=169) started with moderate baseline mobility and experienced moderate decline in mobility over time. The ‘Mid improving” subgroup (n=365) also started with moderate baseline mobility but demonstrated improvement over time. Finally, the “High stable” subgroup (n=60) started with high baseline mobility that remained high during the follow-up period. Subgroups demonstrated similar observation patterns, with comparable numbers of measurements per participant and follow-up duration (Table S5), and similar temporal distribution of observations over follow-up (Figure S2). Finally, the four-class model demonstrated similar overall trajectory patterns (Figure S1).

**Figure 2.**
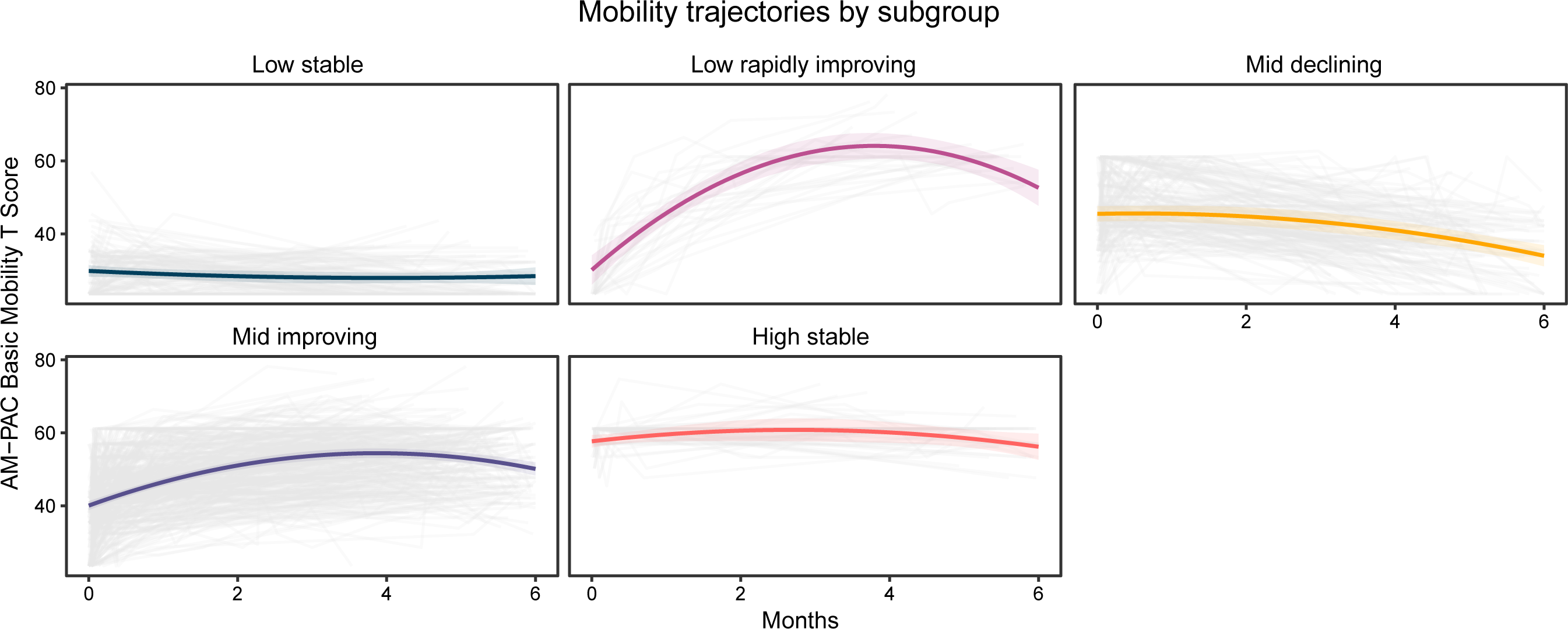
Mobility recovery trajectories for each of the four subgroups. Observed data are shown in grey, while colored lines represent model-estimated trajectories. Shaded ribbons denote 95% confidence intervals. The names of each subgroup were assigned based on the initial functional status and the recovery pattern.

### Subgroup characterization

Each of these subgroups had distinct characteristics as shown in Table 1. Briefly, the “Low stable” subgroup was older, had lower prior levels of independence, higher stroke severity and were more likely to be discharged to a skilled nursing facility compared with other groups. In contrast, the “Low rapidly improving” subgroup tended to be younger, more likely to have been independent prior to hospitalization, received relatively frequent physical therapy during the acute stay, and were likely to be discharged to an inpatient rehabilitation facility. The initial stroke severity and initial ADL and cognitive function were similar to the “Low stable” subgroup, but this group had the lowest proportion of ischemic strokes. The “Mid declining” and “Mid improving” subgroups had higher initial ADL and cognitive function and shorter lengths of stay compared with the “Low stable” and “Low rapidly improving” subgroups. The “Mid declining” subgroup was the less likely to be independent prior to hospitalization, received physical therapy on a smaller proportion of days, and were more likely to be discharged home compared to the “Mid improving” subgroup. The “High stable” subgroup had higher initial ADL and cognitive function, shorter hospital stays, and minimal use of post-acute institutional care compared to all other subgroups. This subgroup also had the highest proportion of ischemic strokes and of individuals who were independent prior to hospitalization. Differences in comorbidities and social factors, including ADI, insurance coverage, and living situation, were less distinct across subgroups. Sensitivity analyses yielded similar patterns of subgroup characteristics when restricting to individuals with high class assignment probability and when weighting by posterior class membership probabilities (Tables S6–S7).

**Table 1.** Characteristics of the five subgroups identified by the growth mixture model.

| Characteristic | Overall<br>N=750* | Low stable<br>N = 127* | Low rapidly<br>improving<br>N = 29* | Mid declining<br>N = 169* | Mid<br>improving<br>N = 365* | High<br>stable<br>N = 60* | p-value<sup>†</sup> |
| --- | --- | --- | --- | --- | --- | --- | --- |
| **Personal Characteristics** |  |  |  |  |  |  |  |
| **Age (years)** | 65 (55, 75) | 71 (63, 79) | 55 (47, 67) | 67 (61, 76) | 63 (52, 75) | 58 (50, 65) | <0.001 |
| **Sex (female)** | 382 (51%) | 69 (54%) | 16 (55%) | 86 (51%) | 187 (51%) | 24 (40%) | 0.5 |
| **Race** |  |  |  |  |  |  | 0.1 |
| Black | 354 (47%) | 59 (46%) | 16 (55%) | 68 (40%) | 179 (49%) | 32 (53%) |  |
| White | 342 (46%) | 60 (47%) | 10 (34%) | 93 (55%) | 153 (42%) | 26 (43%) |  |
| Other | 54 (7.2%) | 8 (6.3%) | 3 (10%) | 8 (4.7%) | 33 (9.0%) | 2 (3.3%) |  |
| **Previous level of function** |  |  |  |  |  |  | <0.001 |
| Independent | 487 (69%) | 60 (52%) | 21 (72%) | 92 (58%) | 270 (78%) | 44 (86%) |  |
| Not Independent | 217 (31%) | 56 (48%) | 8 (28%) | 68 (43%) | 78 (22%) | 7 (14%) |  |
| Unknown | 46 | 11 | 0 | 9 | 17 | 9 |  |
| **History of hypertension** | 676 (90%) | 120 (94%) | 24 (83%) | 157 (93%) | 327 (90%) | 48 (80%) | 0.01 |
| **History of diabetes** | 336 (45%) | 63 (50%) | 12 (41%) | 75 (44%) | 163 (45%) | 23 (38%) | 0.7 |
| **History of hyperlipidemia** | 481 (64%) | 85 (67%) | 17 (59%) | 111 (66%) | 232 (64%) | 36 (60%) | 0.8 |
| **History of depression** | 184 (25%) | 36 (28%) | 5 (17%) | 50 (30%) | 82 (22%) | 11 (18%) | 0.2 |
| **Area Deprivation Index** | 47 (24, 71) | 47 (21, 64) | 42 (19, 74) | 47 (19, 69) | 47 (26, 74) | 46 (25, 71) | 0.8 |
| Unknown | 62 | 7 | 3 | 15 | 32 | 5 |  |
| **Insurance coverage** |  |  |  |  |  |  | 0.005 |
| Medicaid | 60 (8.0%) | 10 (7.9%) | 3 (10%) | 10 (5.9%) | 33 (9.0%) | 4 (6.7%) |  |
| Medicare | 447 (60%) | 82 (65%) | 16 (55%) | 118 (70%) | 209 (57%) | 22 (37%) |  |
| Private | 188 (25%) | 28 (22%) | 9 (31%) | 33 (20%) | 92 (25%) | 26 (43%) |  |
| Other | 55 (7.3%) | 7 (5.5%) | 1 (3.4%) | 8 (4.7%) | 31 (8.5%) | 8 (13%) |  |
| **Living situation** |  |  |  |  |  |  | 0.14 |
| Family | 537 (72%) | 92 (73%) | 27 (93%) | 117 (70%) | 260 (71%) | 41 (72%) |  |

| Characteristic | Overall<br>N=750* | Low stable<br>N = 127* | Low rapidly<br>improving<br>N = 29* | Mid declining<br>N = 169* | Mid<br>improving<br>N = 365* | High<br>stable<br>N = 60* | p-value<sup>†</sup> |
| --- | --- | --- | --- | --- | --- | --- | --- |
| Non-family | 207 (28%) | 34 (27%) | 2 (6.9%) | 51 (30%) | 104 (29%) | 16 (28%) |  |
| Unknown | 6 | 1 | 0 | 1 | 1 | 3 |  |
| **Clinical Characteristics from the Initial Hospitalization** |  |  |  |  |  |  |  |
| **Length of stay (days)** | 8 (5, 14) | 16 (9, 29) | 13 (8, 28) | 6 (4, 11) | 8 (5, 12) | 4 (3, 6) | <0.001 |
| **Initial NIHSS** | 5 (2, 11) | 13 (8,19) | 10 (8,23) | 4 (2,7) | 6 (2,11) | 1 (1,3) | <0.001 |
| Unknown | 335 | 61 | 16 | 82 | 159 | 17 |  |
| **Stroke type (Ischemic)** | 575 (77%) | 92 (72%) | 14 (48%) | 135 (80%) | 282 (78%) | 51 (85%) | 0.001 |
| **Initial AM-PAC Daily Activity T score** | 37 (31, 42) | 31 (29, 33) | 29 (17, 31) | 40 (35, 51) | 36 (31, 40) | 58 (54, 58) | <0.001 |
| Unknown | 26 | 20 | 2 | 3 | 1 | 0 |  |
| **Initial AM-PAC Applied Cognition T score** | 29 (15, 40) | 15 (8, 25) | 17 (11, 26) | 29 (19, 44) | 29 (19, 40) | 62 (44, 62) | <0.001 |
| Unknown | 547 | 91 | 21 | 121 | 267 | 47 |  |
| **Proportion of days visited by physical therapy** | 33 (21, 50) | 34 (22, 56) | 44 (29, 60) | 33 (22, 56) | 40 (29, 58) | 25 (0, 33) | <0.001 |
| **Occupational Therapy involved at acute hospital (yes)** | 5,403 (85%) | 120 (94%) | 27 (93%) | 142 (84%) | 324 (89%) | 44 (73%) | <0.001 |
| **Speech Language Pathology involved at acute hospital (yes)** | 3,994 (63%) | 109 (86%) | 25 (86%) | 107 (63%) | 258 (71%) | 12 (20%) | <0.001 |
| **Discharge location** |  |  |  |  |  |  | <0.001 |
| Inpatient rehabilitation facility | 1,642 (26%) | 40 (31%) | 20 (69%) | 47 (28%) | 202 (55%) | 1 (1.7%) |  |
| Skilled nursing facility | 1,280 (20%) | 75 (59%) | 5 (17%) | 20 (12%) | 45 (12%) | 0 (0%) |  |
| Home | 2,571 (40%) | 12 (9.4%) | 4 (14%) | 102 (60%) | 118 (32%) | 59 (98%) |  |

\* n (%); Median (Q1, Q3)
<sup>†</sup> Pearson's Chi-squared test; Kruskal-Wallis rank sum test

## Discussion

In this study, we identified five distinct trajectories of mobility recovery over the first six months after stroke using longitudinal data derived from the EHR. These subgroups differed not only in baseline mobility but also in their pattern of recovery over time, with some groups demonstrating substantial improvement, others remaining stable, and one experiencing decline. In addition, these trajectories were associated with differences in clinical characteristics, including rehabilitation use during the hospitalization and discharge location. Together, these findings highlight substantial heterogeneity in post-stroke mobility recovery that is not captured by average recovery patterns, which are more commonly used.

Traditionally, stroke recovery is thought of as a process that occurs rapidly initially and then plateaus due to a window of heightened neuroplasticity.^42–44^ In this work, we identified only one group that followed this stereotypical pattern (the “Low rapid improving” subgroup), which was the smallest group in our cohort. This may be related, in part, to how we measured mobility. The AM-PAC Basic Mobility is a self-reported measure of mobility activity, as opposed to a more objective measurement of mobility activity (e.g., gait speed, daily step counts) or motor impairment (e.g., gait parameters). Although this measurement approach may contribute to differences in identified trajectories, patient-reported measures capture the lived experience of mobility and are critical for understanding recovery from the patient’s perspective.^45–48^ Notably, similar trajectory-based analyses of post-stroke upper extremity function that used more objective measures also identified multiple subgroups that do not follow the stereotypical recovery pattern.^15^ This heterogeneity has also been observed using global disability measures, where distinct trajectories characterized by recovery, stability, and persistent disability have been identified following stroke.^49,50^ While different measurement approaches may yield somewhat different subgroup structures, these findings collectively suggest that recovery after stroke is more heterogeneous than implied by the stereotypical model and that group averages may obscure clinically meaningful differences between individuals.

Several of the identified subgroups had similar initial mobility function but exhibited different recovery trajectories. This was most apparent when examining the “Low stable” and “Low rapidly improving” subgroups and the “Mid declining” and “Mid improving” subgroups. These findings align with prior work in upper extremity recovery^15^ and are consistent with patterns observed in clinical practice. Although these subgroup differences suggest that baseline personal and clinical characteristics may contribute to recovery patterns, our analyses were intentionally limited to descriptive characterization of the identified subgroups. We did not evaluate whether individual characteristics were independently associated with trajectory membership. Identification of baseline factors associated with specific recovery trajectories would improve prognostication and represents an important next step.

An important consideration in interpreting the identified subgroups is that they were derived from a subset of stroke survivors with longitudinal mobility assessments available within a single health system. Although 6,366 individuals were admitted with stroke during the study period, only 750 met the requirements for trajectory modeling because repeated mobility measurements were needed to characterize change over time. Most individuals not included in the analysis lacked an AM-PAC Basic Mobility assessment between 91 and 180 days after stroke. Because mobility assessments were obtained as part of routine clinical care rather than a prospective research protocol, the specific reason for missing follow-up data is not know, but may reflect care received outside the health system, death after discharge, reduced healthcare utilization associated with recovery, or other factors. Included participants were younger, more likely to be discharged to an inpatient rehabilitation facility, and more likely to receive rehabilitation services during hospitalization than individuals who were not included. These findings suggest that individuals with ongoing rehabilitation needs may be overrepresented within the analytic cohort. However, among participants with available data, included and excluded individuals appeared broadly similar with respect to stroke type, baseline mobility, activities of daily living function, insurance status, area deprivation, and several other measured characteristics. Therefore, the identified trajectory groups should be interpreted as patterns of mobility recovery among individuals with stroke who remain engaged with the healthcare system to generate longitudinal outcome data available within routine clinical care rather than estimates of recovery trajectories for the broader population of individuals with stroke.

Critical modifiable factors used to characterize our subgroups included rehabilitation use during the acute hospitalization and the discharge location. Individuals in improving subgroups tended to receive physical therapy on a greater proportion of hospital days and were more likely to be discharged to inpatient rehabilitation, whereas those in the high-functioning subgroup had lower exposure to rehabilitation services and were more frequently discharged home. These patterns are consistent with clinical decision-making that aligns rehabilitation intensity with observed or anticipated recovery. However, because rehabilitation exposure and trajectory membership are closely intertwined, these findings should be interpreted cautiously. The observed differences may reflect both appropriate allocation of rehabilitation resources and potential differences in access or responsiveness to care. Although this study was not designed to disentangle these effects, there is a need to understand the causal relationships between these modifiable characteristics and recovery trajectories. Future work should focus on identifying early predictors of trajectory membership, incorporating time-varying measures of rehabilitation exposure, and evaluating whether modifying rehabilitation delivery alters recovery trajectories. In addition, linking trajectory patterns to downstream outcomes such as readmissions, functional independence, and quality of life will be important for determining their clinical significance and guiding targeted intervention development.

This study also has several strengths and limitations. First, EHR data enabled us to characterize recovery in a large sample of individuals with stroke who reflect those receiving clinical care. However, limitations inherent to data collected during routine clinical care must be considered. Missing data were substantial for several variables (e.g., AM-PAC Applied Cognition) due to collection processes within clinical workflows. As a result, some comparisons between included and non-included individuals may be affected by residual uncertainty. In contrast, missingness for the primary outcome measure, AM-PAC Basic Mobility, was minimal among included participants. Additionally, assessment timing and frequency were determined by clinical practice rather than study design. Although growth mixture models can accommodate irregularly timed observations, differences in the number and distribution of assessments may have influenced subgroup identification and stability. Replication across multiple health systems with more complete follow-up and clinical characterization is needed to confirm the observed recovery patterns.

In conclusion, we identified five distinct trajectories of mobility recovery in the first six months after stroke using real-world EHR data. These findings demonstrate that individuals with similar baseline function can follow markedly different recovery paths, highlighting the limitations of relying on average recovery patterns. Improving our ability to identify trajectory membership early in recovery could support more accurate prognostication, better align patient expectations, and guide more targeted rehabilitation strategies, all of which are key goals in optimizing post-stroke care.

## Data Availability

Data is from the electronic medical record and therefore data is not available consistent with the Institution's policy.

## Acknowledgments

We would like to thank Grace Bellinger for extracting the Get With The Guidelines data.

## Sources of Funding

This work was supported by the National Institutes of Health/National Center for Medical Rehabilitation Research (F32HD108835) and the Sheikh Khalifa Stroke Institute.

## Disclosures

None

## Supplemental Material

Supplemental Methods

Tables S1-S7

Figure S1-S2

## Abbreviations

ADI: Area Deprivation Index
ADL: Activities of Daily Living
AM-PAC: Activity Measure for Post-Acute Care
EHR: Electronic Health Record
GMM: Growth Mixture Modeling
GWTG: Get With The Guidelines

## Notes

### Competing Interest Statement

The authors have declared no competing interest.

### Author Declarations

The Johns Hopkins Hospital IRB approved this study (IRB00291279) and granted a waiver of consent.

### Summary of Updates

Added supplemental materials and added clarity on model selection requirements.

