## Supplemental for "Distinct Patterns of Mobility Recovery After Stroke Using Routine Clinical Data"

**Supplemental Materials for “Distinct Patterns of Mobility Recovery After Stroke Using Routine Clinical Data”**

### **Supplemental Methods**

ICD-10 Codes. Stroke diagnoses were identified using ICD-10 codes associated with the hospitalization encounter and billing record. Stroke was defined using ICD-10 code category I60.*-I63.* Comorbid conditions were identified using ICD-10 diagnostic codes and included diabetes (E08.*–E13.*), depression (F06.3*, F31.*–F34.*, F43.2*), hyperlipidemia (E78.*), and hypertension (I10.*–I15.*, I67.4). A diagnosis was considered present if at least one qualifying ICD-10 code was documented in association with the hospitalization encounter and billing record.

### **Table S1**

| **Table S1.** Single‑class model comparisons used to evaluate time functional form and random‑effects structure prior to growth mixture modeling. | | | | | | | | |
| --- | --- | --- | --- | --- | --- | --- | --- | --- |
| **Model** | **Time Term** | **Random intercept** | **Random slope** | **Number of parameters** | **AIC** | **BIC** | **log likelihood** | **p-value** |
| Base linear fixed effects | Linear | No | No | 3 | 26465.9 | 26479.7 | -13229.9 |  |
| Quadratic fixed effects | Quadratic | No | No | 4 | 26379.8 | 26398.2 | -13185.9 | <0.001 |
| Quadratic random intercept | Quadratic | Yes | No | 5 | 25160.1 | 25183.2 | -12575.0 | <0.001 |
| Quadratic random intercept and slope* | Quadratic | Yes | Yes | 7 | 24845.0 | 24877.3 | -12415.5 | <0.001 |
| *These comparisons were conducted prior to class enumeration. Although random slope models improved fit in single‑class models, they produced unstable and poorly separated classes when extended to growth mixture models and were therefore not retained.  *Abbreviations:* AIC- Akaike information criterion; BIC- Bayesian information criterion | | | | | | | | |

### **Table S2**

| **Table S2.** Personal and clinical characteristics of individuals excluded and included in the analytical cohort. | | | | |
| --- | --- | --- | --- | --- |
| **Characteristic** | **Overall**  N = 6,366* | **Excluded**  N = 5,616* | **Included**  N = 750* | **p-value^†^** |
| **Personal Characteristics** | | | | |
| **Age** (years) | 66 (56, 77) | 67 (57, 77) | 65 (55, 75) | 0.002 |
| **Sex** (female) | 3,106 (49%) | 2,724 (49%) | 382 (51%) | 0.4 |
| **Race** |  |  |  | <0.001 |
| Black | 2,562 (40%) | 2,208 (39%) | 354 (47%) |  |
| White | 3,218 (51%) | 2,876 (51%) | 342 (46%) |  |
| Other | 586 (9.2%) | 532 (9.5%) | 54 (7.2%) |  |
| **Previous level of function** |  |  |  | 0.2 |
| Independent | 3,839 (71%) | 3,352 (71%) | 487 (69%) |  |
| Not Independent | 1,566 (29%) | 1,349 (29%) | 217 (31%) |  |
| Unknown | 961 | 915 | 46 |  |
| **History of hypertension** | 5,755 (90%) | 5,079 (90%) | 676 (90%) | 0.8 |
| **History of diabetes** | 2,615 (41%) | 2,279 (41%) | 336 (45%) | 0.03 |
| **History of hyperlipidemia** | 3,821 (60%) | 3,340 (59%) | 481 (64%) | 0.01 |
| **History of depression** | 1,417 (22%) | 1,233 (22%) | 184 (25%) | 0.11 |
| **Area Deprivation Index** | 45 (24, 68) | 45 (24, 67) | 47 (24, 71) | 0.2 |
| Unknown | 645 | 583 | 62 |  |
| **Insurance coverage** |  |  |  | 0.4 |
| Medicaid | 450 (7.1%) | 390 (6.9%) | 60 (8.0%) |  |
| Medicare | 3,681 (58%) | 3,234 (58%) | 447 (60%) |  |
| Private | 1,740 (27%) | 1,552 (28%) | 188 (25%) |  |
| Other | 495 (7.8%) | 440 (7.8%) | 55 (7.3%) |  |
| **Living situation** |  |  |  | 0.7 |
| Family | 4,211 (72%) | 3,674 (72%) | 537 (72%) |  |
| Non-family | 1,670 (28%) | 1,463 (28%) | 207 (28%) |  |
| Unknown | 485 | 479 | 6 |  |
| **Clinical Characteristics from the Initial Hospitalization** | | | | |
| **Length of stay** (days) | 7 (4, 13) | 7 (4, 13) | 8 (5, 14) | 0.9 |
| **Initial NIHSS** | 5 (2, 13) | 5 (2, 13) | 5 (2, 11) | 0.2 |
| Unknown | 2,605 | 2,270 | 335 |  |
| **Stroke type** (Ischemic) | 4,758 (75%) | 4,183 (74%) | 575 (77%) | 0.4 |
| **Initial AM-PAC Daily Activity T score** | 37 (31, 44) | 37 (31, 44) | 37 (31, 42) | 0.6 |
| Unknown | 600 | 574 | 26 |  |
| **Initial AM-PAC Applied Cognition T score** | 27 (15, 40) | 27 (15, 40) | 29 (15, 40) | 0.8 |
| Unknown | 4,685 | 4,138 | 547 |  |
| **Initial AM-PAC Basic Mobility T score** | 38 (26, 48) | 38 (26, 48) | 38 (31, 45) | 0.4 |
| Unknown | 158 | 158 | 0 |  |
| **Proportion of days visited by physical therapy** | 33 (21, 50) | 33 (20, 50) | 38 (25, 55) | <0.001 |
| **Occupational Therapy involved at acute hospital** (yes) | 5,403 (85%) | 4,746 (85%) | 657 (88%) | 0.03 |
| **Speech Language Pathology involved at acute hospital** (yes) | 3,994 (63%) | 3,483 (62%) | 511 (68%) | 0.001 |
| **Discharge location** |  |  |  | <0.001 |
| Inpatient rehabilitation facility | 1,642 (26%) | 1,332 (24%) | 310 (41%) |  |
| Skilled nursing facility | 1,280 (20%) | 1,135 (20%) | 145 (19%) |  |
| Home | 2,571 (40%) | 2,276 (41%) | 295 (39%) |  |
| Other | 459 (7.2%) | 459 (8.2%) | 0 (0%) |  |
| Expired | 414 (6.5%) | 414 (7.4%) | 0 (0%) |  |
| ^1^n (%); Median (Q1, Q3)  ^2^Pearson's Chi-squared test; Welch Two Sample t-test | | | | |

### **Table S3**

| **Table S3.** Fit indices, class sizes, and classification quality metrics for growth mixture models evaluated across two to seven classes. The optimal number of classes was determined by information criteria, the Vuong-Lo-Mendell-Rubin likelihood ratio test, class size (≥5% of sample), and classification quality (mean posterior probability ≥0.7). | | | | | | | |
| --- | --- | --- | --- | --- | --- | --- | --- |
| **Metric** | | **2 Classes** | **3 Classes** | **4 Classes** | **5 Classes** | **6 Classes** | **7 Classes** |
| **Number of parameters estimated** | | 10 | 15 | 20 | 25 | 30 | 35 |
| **AIC** | | 24774.76 | 24724.36 | 24686.26 | 24654.19 | 24625.93 | 24602.42 |
| **BIC** | | 24820.96 | 24793.66 | 24778.66 | 24769.69 | 24764.53 | 24764.12 |
| **Sample size adjusted BIC** | | 24789.21 | 24746.03 | 24715.16 | 24690.31 | 24669.27 | 24652.98 |
| **Entropy** | | 0.64 | 0.67 | 0.62 | 0.73 | 0.7 | 0.65 |
| **Likelihood Ratio Test p value** | |  | <0.001 | <0.001 | <0.001 | <0.001 | <0.001 |
| **Class size (n)** | **Class 1** | 310 (41.3%) | 289 (38.5%) | 100 (13.3%) | 127 (16.9%) | 128 (17.1%) | 124 (16.5%) |
|  | **Class 2** | 440 (58.7%) | 130 (17.3%) | 131 (17.5%) | 365 (48.7%) | 33 (4.4%) | 215 (28.7%) |
|  | **Class 3** |  | 331 (44.1%) | 188 (25.1%) | 29 (3.9%) | 124 (16.5%) | 40 (5.3%) |
|  | **Class 4** |  |  | 331 (44.1%) | 59 (7.9%) | 366 (48.8%) | 38 (5.1%) |
|  | **Class 5** |  |  |  | 170 (22.7%) | 32 (4.3%) | 77 (10.3%) |
|  | **Class 6** |  |  |  |  | 67 (8.9%) | 135 (18%) |
|  | **Class 7** |  |  |  |  |  | 121 (16.1%) |
| **Mean probability of class assignment** | **Class 1** | 0.89 | 0.84 | 0.76 | 0.82 | 0.76 | 0.74 |
|  | **Class 2** | 0.89 | 0.78 | 0.80 | 0.85 | 0.72 | 0.71 |
|  | **Class 3** |  | 0.88 | 0.81 | 0.81 | 0.81 | 0.73 |
|  | **Class 4** |  |  | 0.76 | 0.78 | 0.81 | 0.69 |
|  | **Class 5** |  |  |  | 0.81 | 0.81 | 0.78 |
|  | **Class 6** |  |  |  |  | 0.75 | 0.66 |
|  | **Class 7** |  |  |  |  |  | 0.82 |

### **Table S4**

| **Table S4.** Parameter estimates for the selected 5‑class growth mixture model. Values presented are estimated coefficients with standard errors. | | | | | |
| --- | --- | --- | --- | --- | --- |
| **Parameter** | **Class 1**  N = 127 | **Class 3**  N = 29 | **Class 5**  N = 169 | **Class 2**  N = 365 | **Class 4**  N = 60 |
| Assigned Name | Low stable | Low rapidly improving | Mid declining | Mid improving | High stable |
| Estimated baseline AM-PAC Mobility T score (Day 0) | 29.8 (0.8) | 30.1 (2.0) | 45.5 (1.1) | 40.1 (0.6) | 57.7 (0.9) |
| Estimated linear change in AM-PAC Mobility T score (per month) | -1.0 (0.7) | 17.9 (1.9) | 0.4 (0.7) | 7.4 (0.5) | 2.3 (1.3) |
| Estimated quadratic change in AM-PAC Mobility T score (per month^2^) | 0.1 (0.1) | -2.4 (0.3) | -0.4 (0.1) | -1.0 (0.08) | -0.4 (0.2) |

### **Table S5**

| **Table S5.** Number and timing of measurements per participant across classes | | | | | | | |
| --- | --- | --- | --- | --- | --- | --- | --- |
| **Subgroup** | **Observations per person*** | **Day of first observation (days post stroke)*** | | **Day of last observation (days post stroke)*** | | | **Time between first and last measurement (days)*** |
| **1 (Low stable)** | 4 (3-4) | 1 (0.5, 1) | | 137 (109.5, 168.5) | | | 136 (108.5, 168) |
| **3 (Low rapidly improving)** | 5 (4-7) | 1 (1, 1) | | 153 (119, 164) | | | 152 (118, 162) |
| **5 (Mid declining)** | 4 (3-5) | 1 (1, 1) | | 141 (115, 165) | | | 140 (115, 164) |
| **2 (Mid improving)** | 4 (3-6) | 1 (1, 1) | | 141 (118, 165) | | | 141 (117, 164) |
| **4 (High stable)** | 4 (3-5) | 1 (1, 1) | | 144 (120, 171.5) | | | 143 (118, 170) |
| * Median (Q1-Q3) |  | |  | |  |  | |

### **Table S6**

| **Table S6.** Characteristics of participants with high probability class assignment (≥0.7). This was conducted as a sensitivity analysis since class assignment is unknown and probabilistic. | | | | | | | |
| --- | --- | --- | --- | --- | --- | --- | --- |
| **Characteristic** | **Overall** N=589* | **Low stable**  N = 95* | **Low rapidly improving**  N = 23* | **Mid declining**  N = 124* | **Mid improving**  N = 302* | **High stable**  N = 45* | **p-value^†^** |
| **Personal Characteristics** | | | | | | | |
| **Age** (years) | 66 (56, 75) | 72 (64, 79) | 57 (47, 68) | 66 (61, 75) | 64 (53, 75) | 58 (49, 65) | <0.001 |
| **Sex** (female) | 298 (51%) | 53 (56%) | 14 (61%) | 58 (47%) | 155 (51%) | 18 (40%) | 0.3 |
| **Race** |  |  |  |  |  |  | 0.3 |
| Black | 276 (47%) | 46 (48%) | 12 (52%) | 46 (37%) | 147 (49%) | 25 (56%) |  |
| White | 272 (46%) | 43 (45%) | 10 (43%) | 71 (57%) | 130 (43%) | 18 (40%) |  |
| Other | 41 (7.0%) | 6 (6.3%) | 1 (4.3%) | 7 (5.6%) | 25 (8.3%) | 2 (4.4%) |  |
| **Previous level of function** |  |  |  |  |  |  | <0.001 |
| Independent | 387 (70%) | 46 (53%) | 17 (74%) | 68 (58%) | 223 (77%) | 33 (87%) |  |
| Not Independent | 168 (30%) | 41 (47%) | 6 (26%) | 49 (42%) | 67 (23%) | 5 (13%) |  |
| Unknown | 34 | 8 | 0 | 7 | 12 | 7 |  |
| **History of hypertension** | 534 (91%) | 90 (95%) | 20 (87%) | 116 (94%) | 273 (90%) | 35 (78%) | 0.023 |
| **History of diabetes** | 265 (45%) | 49 (52%) | 10 (43%) | 58 (47%) | 131 (43%) | 17 (38%) | 0.5 |
| **History of hyperlipidemia** | 384 (65%) | 63 (66%) | 13 (57%) | 83 (67%) | 196 (65%) | 29 (64%) | >0.9 |
| **History of depression** | 146 (25%) | 21 (22%) | 5 (22%) | 39 (31%) | 72 (24%) | 9 (20%) | 0.4 |
| **Area Deprivation Index** | 46 (22, 71) | 50 (23, 66) | 39 (19, 59) | 44 (17, 66) | 45 (23, 74) | 46 (23, 73) | 0.6 |
| Unknown | 50 | 5 | 2 | 13 | 25 | 5 |  |
| **Insurance coverage** |  |  |  |  |  |  | 0.1 |
| Medicaid | 47 (8.0%) | 6 (6.3%) | 2 (8.7%) | 8 (6.5%) | 27 (8.9%) | 4 (8.9%) |  |
| Medicare | 343 (58%) | 60 (63%) | 14 (61%) | 82 (66%) | 172 (57%) | 15 (33%) |  |
| Private | 156 (26%) | 23 (24%) | 6 (26%) | 28 (23%) | 79 (26%) | 20 (44%) |  |
| Other | 43 (7.3%) | 6 (6.3%) | 1 (4.3%) | 6 (4.8%) | 24 (7.9%) | 6 (13%) |  |
| **Living situation** |  |  |  |  |  |  | 0.3 |
| Family | 428 (73%) | 66 (70%) | 21 (91%) | 92 (75%) | 216 (72%) | 33 (77%) |  |
| Non-family | 157 (27%) | 28 (30%) | 2 (8.7%) | 31 (25%) | 86 (28%) | 10 (23%) |  |
| Unknown | 4 | 1 | 0 | 1 | 0 | 2 |  |
| **Clinical Characteristics from the Initial Hospitalization** | | | | | | | |
| **Length of stay** (days) | 8 (5, 14) | 16 (9, 29) | 13 (8, 28) | 6 (4, 12) | 8 (5, 13) | 4 (3, 6) | <0.001 |
| **Initial NIHSS** | 6 (2,12) | 14 (9, 19) | 10 (8, 23) | 4 (2, 7) | 6 (3,12) | 1 (0,2) | <0.001 |
| Unknown | 258 | 43 | 13 | 57 | 130 | 15 |  |
| **Stroke type** (Ischemic) | 450 (76%) | 67 (71%) | 9 (39%) | 101 (81%) | 235 (78%) | 38 (84%) | <0.001 |
| **Initial AM-PAC Daily Activity T score** | 36 (31, 42) | 31 (29, 31) | 31 (17, 31) | 40 (36, 51) | 36 (31, 40) | 58 (58, 58) | <0.001 |
| Unknown | 24 | 19 | 2 | 3 | 0 | 0 |  |
| **Initial AM-PAC Applied Cognition T score** | 29 (19, 40) | 15 (8, 25) | 19 (15, 29) | 31 (22, 48) | 29 (19, 40) | 62 (53, 62) | <0.001 |
| Unknown | 428 | 69 | 16 | 90 | 219 | 34 |  |
| **Proportion of days visited by physical therapy** | 37 (25, 54) | 31 (22, 56) | 43 (24, 59) | 33 (21, 53) | 40 (29, 57) | 20 (0, 33) | <0.001 |
| **Occupational Therapy involved at acute hospital** (yes) | 519 (88%) | 91 (96%) | 21 (91%) | 103 (83%) | 272 (90%) | 32 (71%) | <0.001 |
| **Speech Language Pathology involved at acute hospital** (yes) | 408 (69%) | 84 (88%) | 20 (87%) | 81 (65%) | 216 (72%) | 7 (16%) | <0.001 |
| **Discharge location** |  |  |  |  |  |  | <0.001 |
| Inpatient rehabilitation facility | 254 (43%) | 29 (31%) | 16 (70%) | 34 (27%) | 174 (58%) | 1 (2.2%) |  |
| Skilled nursing facility | 115 (20%) | 60 (63%) | 4 (17%) | 13 (10%) | 38 (13%) | 0 (0%) |  |
| Home | 220 (37%) | 6 (6.3%) | 3 (13%) | 77 (62%) | 90 (30%) | 44 (98%) |  |
| * n (%); Median (Q1, Q3)  **^†^** Pearson's Chi-squared test; Kruskal-Wallis rank sum test | | | | | | | |

### **Table S7**

| **Table S7.** Probability weighted analysis. Everyone is included in each subgroup with their values weighted according to the posterior probability of membership in that subgroup. | | | | | |
| --- | --- | --- | --- | --- | --- |
| **Characteristic** | **Low stable**^*^ | **Low rapidly improving**^*^ | **Mid declining**^*^ | **Mid improving**^*^ | **High stable**^*^ |
| **Personal Characteristics** | | | | | |
| **Age** (years) | 71.4 (63.0, 79.0) | 56.9 (45.9, 67.3) | 66.7 (60.0, 76.0) | 64.0 (53.0, 75.0) | 58.0 (50.3, 66.9) |
| **Sex** (female) | 63.7 (55.0%) | 20.4 (52.5%) | 93.7 (50.8%) | 178.5 (51.2%) | 25.7 (41.3%) |
| **Race** |  |  |  |  |  |
| Black | 54.7 (47.3%) | 20.6 (53.1%) | 77.5 (42.0%) | 168.4 (48.3%) | 32.8 (52.6%) |
| White | 53.0 (45.8%) | 14.4 (37.2%) | 97.2 (52.7%) | 150.7 (43.2%) | 26.7 (42.8%) |
| Other | 8.0 (6.9%) | 3.8 (9.7%) | 9.7 (5.3%) | 29.6 (8.5%) | 2.9 (4.6%) |
| **Previous level of function** |  |  |  |  |  |
| Independent | 56.2 (53.3%) | 29.0 (77.3%) | 103.1 (59.3%) | 252.0 (75.6%) | 46.6 (86.7%) |
| Not Independent | 49.2 (46.7%) | 8.5 (22.7%) | 70.6 (40.7%) | 81.5 (24.4%) | 7.2 (13.3%) |
| Unknown | 46 | 46 | 46 | 46 | 46 |
| **History of hypertension** | 109.3 (94.5%) | 32.7 (84.3%) | 169.9 (92.1%) | 314.1 (90.0%) | 50.0 (80.3%) |
| **History of diabetes** | 58.9 (50.9%) | 15.2 (39.2%) | 84.8 (46.0%) | 152.9 (43.8%) | 24.2 (38.8%) |
| **History of hyperlipidemia** | 76.7 (66.3%) | 21.1 (54.4%) | 124.0 (67.3%) | 221.2 (63.4%) | 37.9 (60.9%) |
| **History of depression** | 30.3 (26.2%) | 6.7 (17.2%) | 53.4 (29.0%) | 80.9 (23.2%) | 12.7 (20.4%) |
| **Area Deprivation Index** | 50.0 (23.0, 65.4) | 43.7 (24.0, 73.2) | 47.3 (24.9, 69.0) | 47.0 (25.0, 74.0) | 45.6 (26.8, 69.9) |
| Unknown | 62 | 62 | 62 | 62 | 62 |
| **Insurance coverage** |  |  |  |  |  |
| Medicaid | 8.6 (7.4%) | 4.0 (10.4%) | 11.3 (6.1%) | 31.0 (8.9%) | 5.1 (8.2%) |
| Medicare | 76.5 (66.1%) | 20.8 (53.7%) | 125.2 (67.9%) | 200.2 (57.4%) | 24.2 (38.9%) |
| Private | 24.2 (20.9%) | 10.9 (28.1%) | 37.4 (20.3%) | 89.8 (25.7%) | 25.7 (41.2%) |
| Other | 6.5 (5.6%) | 3.0 (7.8%) | 10.5 (5.7%) | 27.8 (8.0%) | 7.3 (11.7%) |
| **Living situation** |  |  |  |  |  |
| Family | 82.1 (71.6%) | 32.4 (83.5%) | 128.8 (70.4%) | 250.8 (72.1%) | 42.9 (71.8%) |
| Non-family | 32.5 (28.4%) | 6.4 (16.5%) | 54.1 (29.6%) | 97.2 (27.9%) | 16.9 (28.2%) |
| Unknown | 6 | 6 | 6 | 6 | 6 |
| **Clinical Characteristics from the Initial Hospitalization** | | | | | |
| **Length of stay** (days) | 16.0 (9.0, 29.0) | 12.9 (8.0, 25.7) | 6.7 (4.0, 12.0) | 7.9 (5.0, 13.0) | 4.0 (3.0, 7.0) |
| **Initial NIHSS** | 13.0 (8.7, 19.0) | 10.0 (8.0, 20.9) | 4.0 (2.0, 8.0) | 5.0 (2.0, 11.0) | 1.0 (1.0, 3.0) |
| Unknown | 335 | 335 | 335 | 335 | 335 |
| **Stroke type** (Ischemic) | 82.1 (70.9%) | 21.5 (55.5%) | 149.5 (81.1%) | 269.9 (77.4%) | 52.0 (83.5%) |
| **Initial AM-PAC Daily Activity T score** | 30.6 (29.0, 32.4) | 30.6 (29.0, 33.4) | 38.7 (33.4, 47.1) | 36.0 (30.6, 40.2) | 57.5 (47.1, 57.5) |
| Unknown | 26 | 26 | 26 | 26 | 26 |
| **Initial AM-PAC Applied Cognition T score** | 15.2 (8.8, 26.7) | 22.6 (15.2, 31.4) | 29.7 (20.2, 44.3) | 28.8 (19.3, 39.0) | 62.2 (41.5, 62.2) |
| Unknown | 547 | 547 | 547 | 547 | 547 |
| **Proportion of days visited by physical therapy** | 33.6 (23.1, 55.6) | 48.4 (29.6, 61.9) | 33.3 (23.0, 53.3) | 40.0 (28.6, 58.3) | 25.5 (0.0, 39.9) |
| **Occupational Therapy involved at acute hospital** (yes) | 109.7 (94.8%) | 35.4 (91.2%) | 156.3 (84.8%) | 310.5 (89.0%) | 45.1 (72.4%) |
| **Speech Language Pathology involved at acute hospital** (yes) | 99.7 (86.2%) | 33.8 (87.1%) | 117.2 (63.5%) | 243.5 (69.8%) | 16.8 (27.0%) |
| **Discharge location** |  |  |  |  |  |
| Inpatient rehabilitation facility | 38.3 (33.1%) | 27.4 (70.7%) | 55.0 (29.8%) | 187.4 (53.7%) | 1.9 (3.1%) |
| Skilled nursing facility | 65.6 (56.7%) | 5.8 (15.0%) | 27.8 (15.1%) | 45.2 (13.0%) | 0.5 (0.8%) |
| Home | 11.8 (10.2%) | 5.5 (14.3%) | 101.6 (55.1%) | 116.2 (33.3%) | 59.9 (96.2%) |
| ^*^ Weighted median (Q1, Q3); weighted proportions n (%) | | | | | |

### **Figure S1**

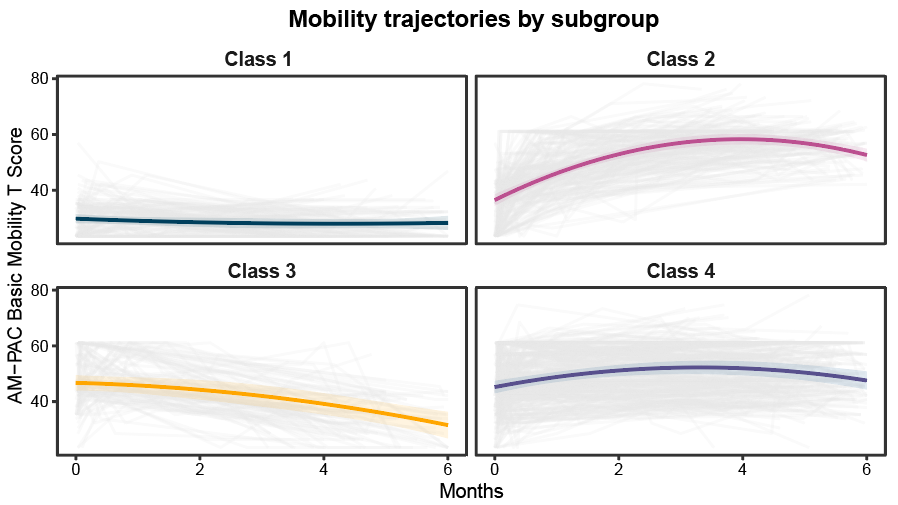

**Figure S1.** Subgroups from the 4-class solution. Subgroups are similar to those identified in the 5-class solution; however, the 5-class solution had lower information criteria and higher entropy indicating a better model fit.

### **Figure S2.**

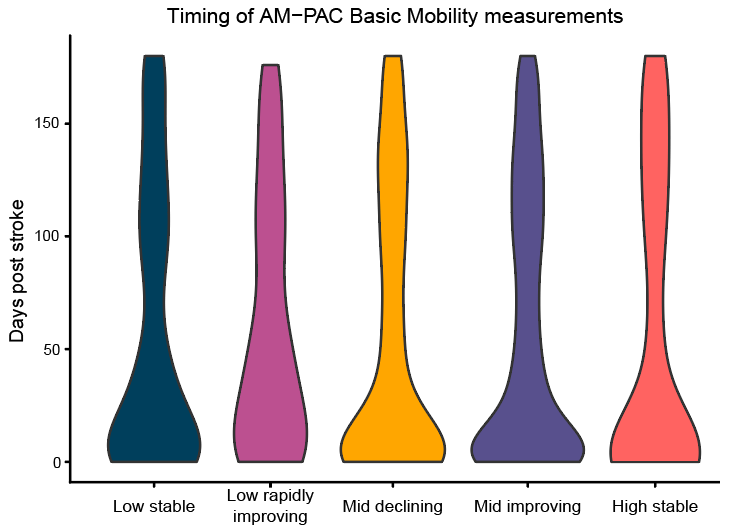

**Figure S2.** Timing of measurements by subgroup.
